# Nationwide Analysis of Etiologies and Risk Factors for Early Readmissions After Hospitalization for Endophthalmitis in the United States

**DOI:** 10.1101/2025.09.08.25335321

**Authors:** Mukund Garg, Sonali Gupta

## Abstract

**Purpose:** To study the patterns of 90-day readmissions after hospitalization for endophthalmitis.

**Methods:** Using the Nationwide Readmissions Database, hospitalizations due to endophthalmitis were identified. Primary outcome was readmission within 90 days. Survival analysis was performed, and predictors of readmission were analyzed using multivariable Cox proportional hazards regression.

**Results:** 1865 patients met the inclusion criteria (mean age 58.3 years, female 47%). Of these, 440 (24%) patients were readmitted within 90 days. Most common reasons for readmissions were sepsis (14%), endophthalmitis (10%), acute renal failure (3%), pneumonia (3%), and corneal ulcer (3%). Heart failure (HR: 1.8, 95% CI: 1.3-2.6), liver cirrhosis (HR: 2.3, 95% CI: 1.4-3.7), infective endocarditis (HR: 2.5, 95% CI: 1.4-4.3), and urinary tract infection (HR: 1.8, 95% CI: 1.1-2.9) were independently associated with a higher risk of readmission.

**Conclusion:** Nearly one in four patients hospitalized due to endophthalmitis get readmitted within 90 days, and most readmissions are due to recurrent endophthalmitis, other ocular complications, and systemic infections.

## Introduction

Endophthalmitis is a devastating intraocular infection and vision-threatening emergency that requires urgent management. Despite advances in antimicrobial therapy, intravitreal injection techniques, and vitreoretinal surgery, endophthalmitis continues to be associated with poor visual outcomes and high health care utilization (1,2). Prior research has largely focused on microbiology, treatment strategies, and visual prognosis (3), but hospital readmissions in this population have not been well characterized.

Hospital readmissions are increasingly viewed as a quality metric and a driver of health care costs (4,5), making them a critical area of investigation in ophthalmology. Identifying predictors of readmission could highlight vulnerable patient groups and inform strategies for post-discharge monitoring and multidisciplinary care. In this study, we used a large, nationally representative database to characterize the incidence, causes, and risk factors for 90-day readmissions following hospitalization for endophthalmitis.

## Methods

We conducted a retrospective cohort study using the Nationwide Readmissions Database (NRD) for 2016–2018 and 2021. The NRD is sponsored by the Agency for Healthcare Research and Quality and captures hospital discharge data from community hospitals across 28 U.S. states, representing approximately 60% of the U.S. population. The NRD allows for longitudinal tracking of patients across hospitalizations within a single calendar year.

Patients with a primary diagnosis of endophthalmitis were identified using International Classification of Diseases, Tenth Revision (ICD-10) codes. We excluded patients who died during the index admission, those discharged in October–December (to ensure a 90-day follow-up window), and non-residents of the state in which they were hospitalized. After applying these exclusions, the final analytic cohort included 1,865 patients.

The primary outcome was all-cause readmission within 90 days of discharge. Secondary outcomes included the most common principal diagnoses associated with readmissions. Covariates included patient demographics, median income quartile by zip code, comorbidities, infection-related conditions, and hospital characteristics such as teaching status and bed size. Comorbidities of interest included diabetes, chronic kidney disease, cirrhosis, congestive heart failure, chronic obstructive pulmonary disease, malignancy, urinary tract infection, pneumonia, and infective endocarditis.

Comparisons between patients with and without readmissions were performed using chi-square tests for categorical variables and t-tests for continuous variables. Cox proportional hazards regression was used to assess predictors of 90-day readmission. Variables with p<0.10 in univariate analysis were included in the multivariable model, and hazard ratios with 95% confidence intervals were reported. Analyses accounted for discharge-level weights to generate nationally representative estimates.

## Results

A total of 1,865 patients were hospitalized with a primary diagnosis of endophthalmitis during the study period. The mean age was 58.3 years (SD 20.2), and 47% were female. Of these, 440 patients (23.6%) were readmitted within 90 days. Baseline characteristics comparing readmitted and non-readmitted patients are presented in **Table 1**. Patients who were readmitted were more likely to have diabetes, chronic kidney disease, cirrhosis, congestive heart failure, chronic obstructive pulmonary disease, and malignancy. They were also more likely to be discharged to a non-home setting and incurred higher hospital charges.

**Table 1.**
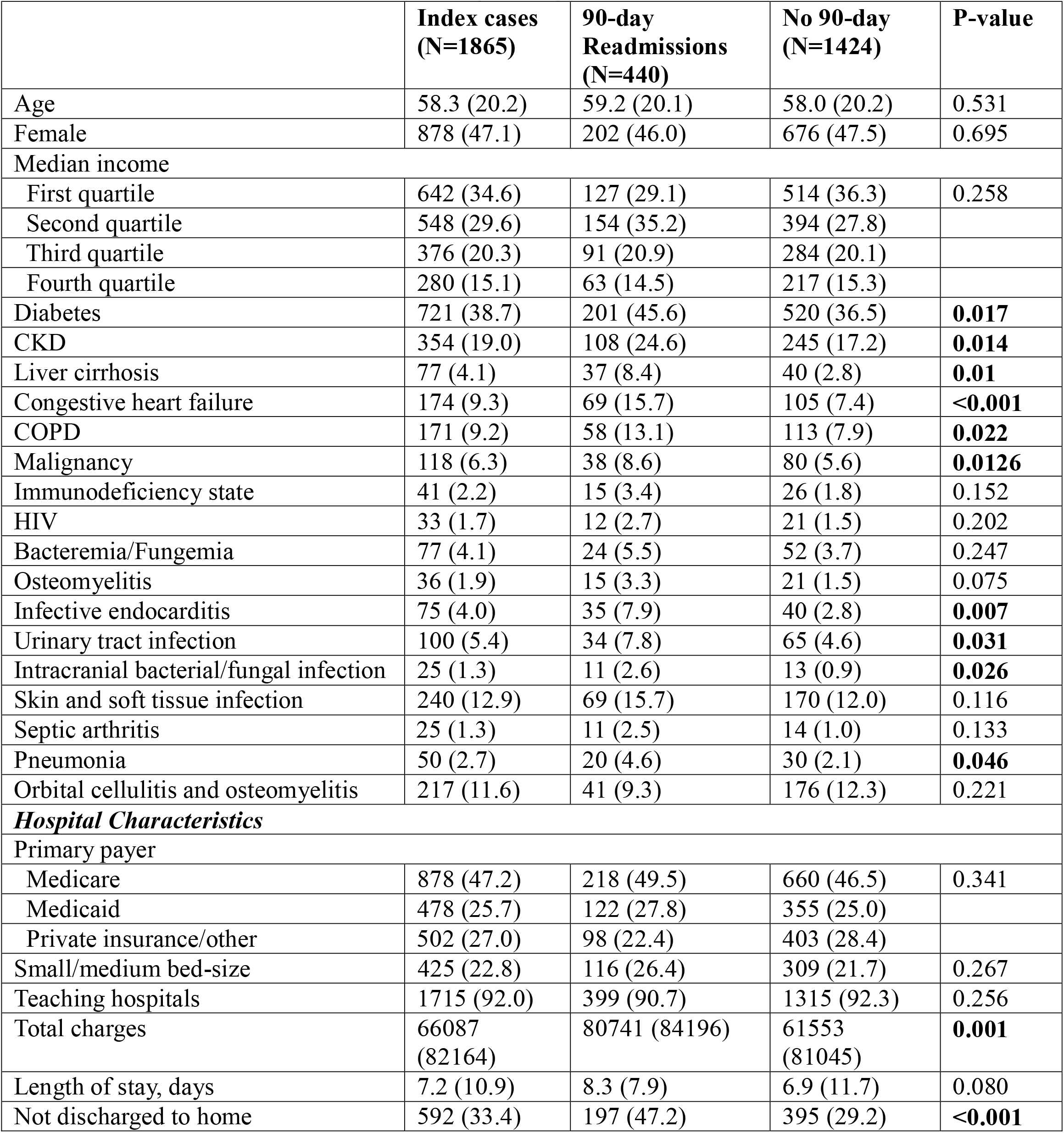
Baseline characteristics stratified by 90-day readmissions.

The most frequent causes of readmission are shown in **Table 2**. Sepsis accounted for 13.9% of readmissions, followed by recurrent endophthalmitis (10.0%), acute kidney failure (3.2%), pneumonia (3.2%), and corneal ulcer (2.7%). Infective endocarditis was the principal diagnosis in 1.8% of readmissions.

**Table 2.** Most common etiologies of 90-day Readmissions (N=440)

| **Table 2. Most common etiologies of 90-day Readmissions**<br>**(N=440)** |  |
| --- | --- |
| Sepsis | 61 (13.9) |
| Endophthalmitis | 44 (10.0) |
| Acute kidney failure | 14 (3.2) |
| Pneumonia | 14 (3.2) |
| Corneal ulcer | 12 (2.7) |
| Infective endocarditis | 8 (1.8) |

On univariate analysis, several comorbidities including diabetes, chronic kidney disease, cirrhosis, congestive heart failure, chronic obstructive pulmonary disease, urinary tract infection, pneumonia, and infective endocarditis were associated with an increased risk of 90-day readmission. Results of the multivariable Cox regression analysis are presented in **Table 3**. Independent predictors of readmission included liver cirrhosis (HR 2.29, 95% CI 1.42–3.69, p=0.001), congestive heart failure (HR 1.81, 95% CI 1.26–2.60, p=0.001), infective endocarditis (HR 2.45, 95% CI 1.40–4.28, p=0.002), and urinary tract infection (HR 1.75, 95% CI 1.06–2.88, p=0.028). The Kaplan–Meier curve of readmission-free survival is displayed in **Figure 1**, which shows the steepest decline in survival within the first 30 days post-discharge.

**Fig. 1.**
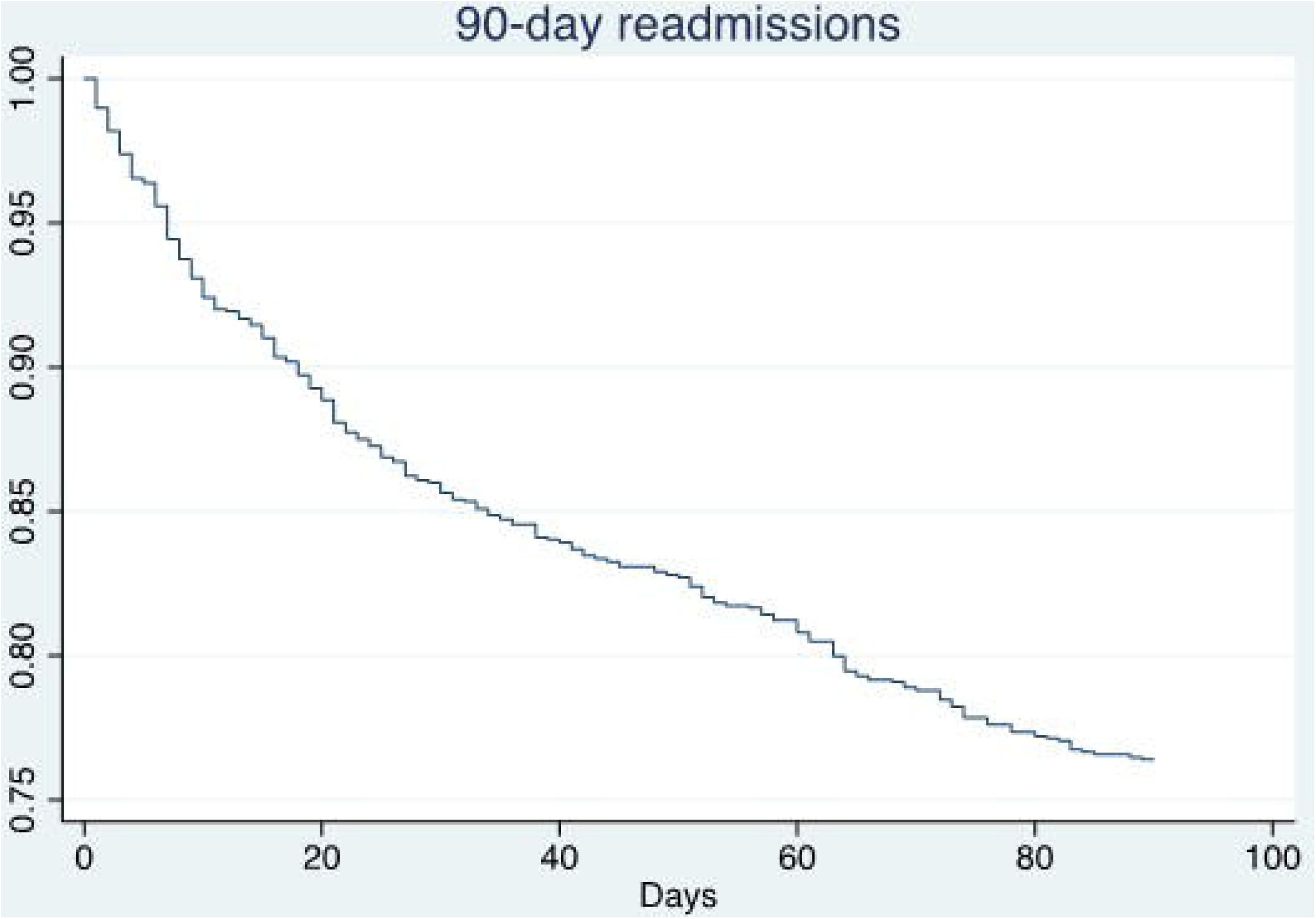

**Table 3.** Predictors of 90-day Readmissions.

| **Table 3. Predictors of 90-day Readmissions** |  |  |
| --- | --- | --- |
|  | **Univariate Analysis** |  |
| **Variables** | **HR (95% CI)** | **P-value** |
| Age, per 10-year increase | 1.03 (0.95-1.11) | 0.442 |
| Women | 0.96 (0.74-1.24) | 0.753 |
| Median income (>50<sup>th</sup> centile) | 0.99 (0.72-1.36) | 0.953 |
| Diabetes | 1.39 (1.06-1.83) | **0.016** |
| CKD | 1.49 (1.09-2.02) | **0.011** |
| Liver cirrhosis | 2.39 (1.44-3.96) | **0.001** |
| Congestive heart failure | 1.96 (1.38-2.78) | **<0.001** |
| COPD | 1.52 (1.05-2.19) | **0.026** |
| Malignancy | 1.48 (0.93-2.38) | 0.101 |
| Immunodeficiency state | 1.94 (0.87-4.30) | 0.103 |
| HIV | 1.61 (0.77-3.38) | 0.209 |
| Bacteremia/Fungemia | 1.41 (0.80-2.50) | 0.235 |
| Osteomyelitis | 2.07 (1.00-4.30) | 0.050 |
| Infective endocarditis | 2.54 (1.37-4.69) | **0.003** |
| Urinary tract infection | 1.63 (1.06-2.51) | **0.027** |
| Intracranial bacterial/fungal infection | 2.38 (1.16-4.88) | **0.018** |
| Skin and soft tissue infection | 1.32 (0.94-1.85) | 0.106 |
| Septic arthritis | 1.92 (0.84-4.35) | 0.120 |
| Pneumonia | 1.93 (1.03-3.59) | **0.039** |
| Orbital cellulitis and osteomyelitis | 0.80 (0.50-1.23) | 0.335 |
| Private/other insurance | 0.75 (0.52-1.07) | 0.115 |
| Small/medium bed-size | 1.25 (0.85-1.85) | 0.262 |
| Teaching hospitals | 0.84 (0.61-1.14) | 0.271 |

## Discussion

This large, nationwide study demonstrates that nearly one in four patients hospitalized with endophthalmitis are readmitted within 90 days. The distribution of readmission causes (Table 2) indicates that systemic infections such as sepsis and pneumonia were more common than recurrent ocular infections, underscoring the vulnerability of this population to systemic illness. The Kaplan–Meier curve (Figure 1) further illustrates that the highest risk period is within the first month after discharge, suggesting a critical window for follow-up and preventive strategies.

Our multivariable analysis (Table 3) identified cirrhosis, congestive heart failure, infective endocarditis, and urinary tract infection as independent predictors of readmission. These comorbidities are known to compromise host defenses and may complicate recovery from ocular infections. Previous literature has highlighted the interplay between systemic health and ocular outcomes (1,3), and our findings reinforce the importance of multidisciplinary care. Patients with these risk factors may benefit from enhanced discharge planning, early outpatient follow-up, and coordination between ophthalmologists, primary care physicians, and infectious disease specialists.

Strengths of this study include the use of a large, nationally representative dataset and robust multivariable modeling. Limitations include reliance on administrative coding, lack of ophthalmic details such as microbiological data and visual outcomes, and inability to capture readmissions across state lines or outpatient encounters.

## Conclusion

In this nationwide analysis, nearly one quarter of patients hospitalized with endophthalmitis were readmitted within 90 days. While recurrent ocular infection was a common cause, systemic infections such as sepsis were the most frequent readmission diagnoses. Cirrhosis, congestive heart failure, infective endocarditis, and urinary tract infection were independent predictors of readmission. Recognizing these risk factors at the time of discharge may guide targeted follow-up strategies, multidisciplinary management, and prevention efforts to reduce readmissions and improve outcomes.

## Data Availability

Data in the study were obtained from the Natiowide Readmissions Database that is publicly available through the HCUP website: https://hcup-us.ahrq.gov/nrdoverview.jsp

## Declarations

### Author Approval

All authors have seen and approved the manuscript.

### Competing Interests

None

### Data Availability

We obtained data from the Nationwide Readmissions Database that is a publicly available database that is available through the HCUP website.

### Funding

None

## Notes

### Competing Interest Statement

The authors have declared no competing interest.

### Funding Statement

This study did not receive any funding

